# Survival benefit of adjuvant therapy following neoadjuvant therapy in patients with resected esophageal cancer: a retrospective cohort study

**DOI:** 10.1101/2024.05.23.24307798

**Authors:** Weiyi Jia, Chao Li, Can Liu, Renwang Hu

**Affiliations:** Department of Gastrointestinal Surgery, Henan Provincial People’s Hospital, Zhengzhou, Henan, China; Department of Science & Education, Zhengzhou Central Hospital Affiliated to Zhengzhou University, Zhengzhou 450007, PR China; Department of Radiology, Henan Provincial People’s Hospital, Zhengzhou, Henan, China

**Author notes:** Correspondence: Renwang Hu, MD. **Address:** 7 Weiwu Road. Zhengzhou, Henan 450000, China. **E-mail:**.

**Keywords:** esophageal cancer, neoadjuvant therapy, adjuvant therapy, survival analysis

## Abstract

**Background:** There is controversy about the benefit of administering adjuvant therapy to esophageal cancer (EC) patients after preoperative neoadjuvant therapy and surgical treatment. This study aims to investigate the impact of postoperative adjuvant therapy in EC patients with neoadjuvant therapy and surgery.

**Materials and methods:** The study included EC patients diagnosed from 2007 to 2020 in the Surveillance, Epidemiology, and End Results (SEER) database. Patients who received neoadjuvant therapy (NCRT) were defined as those who underwent neoadjuvant chemotherapy or neoadjuvant radiotherapy before surgery, while patients who received adjuvant therapy (ACRT) were defined as those who underwent adjuvant chemotherapy or adjuvant radiotherapy after surgery. Propensity score matching (PSM) method was employed to establish matched cohorts, and Kaplan-Meier analysis, COX regression model, and Fine-Gray model were used for survival analysis.

**Results:** The study included a total of 5805 EC patients, with 837 (14.4%) in the ACRT group and 4968 (85.4%) in the no-ACRT group. After PSM, a cohort of 1660 patients who received NCRT was enrolled for analysis, with 830 patients in each group. Kaplan-Meier analysis revealed no significant differences between the two groups in terms of median overall survival (OS) (34.0 vs. 36.0 months, P = 0.89) or cancer-specific survival (CSS) (40.0 vs. 49.0 months, P = 0.16). Multivariate Cox models and Fine-Gray models indicated that ACRT was not a predictive factor for OS or CSS (P > 0.05). Subgroup analysis for CSS suggested a protective effect of ACRT in the N2 (Cox model: HR = 0.640, P = 0.090; Fine-Gray model: HR = 0.636, P = 0.081) and the N3 subgroup (Cox model: HR = 0.302, P = 0.018; Fine-Gray model: HR = 0.306, P = 0.034).

**Conclusions:** Only for esophageal cancer patients with a more advanced N stage, postoperative adjuvant therapy after completing neoadjuvant therapy and curative surgical treatment may be beneficial.

## Introduction

Esophageal cancer (EC) is an important health concern worldwide, ranking seventh in incidence and sixth in cancer-related mortality globally [1]. Surgical treatment remains the foremost curative approach for esophageal cancer, yet the prognosis with surgery alone often falls short of satisfaction [2]. Consequently, investigations into the benefits of postoperative adjuvant therapy and preoperative neoadjuvant therapy have been undertaken. Postoperative adjuvant therapy was believed to clear residual tumors that are difficult to detect, while neoadjuvant therapy was thought to address early micro-metastases, downsize the primary tumor, and improve the R0 resection rate for esophageal cancer [3–7]. In numerous studies, both adjuvant therapy and neoadjuvant therapy have demonstrated substantial survival benefits [8–12].

Currently, radical surgery following neoadjuvant therapy has become the standard treatment approach for locally advanced esophageal cancer [3, 13, 14]. However, the efficacy of postoperative adjuvant therapy in EC patients who have undergone neoadjuvant therapy and radical surgery remains a subject of controversy [14–16]. Research exploring this issue was quite limited, and the results for this matter would play an important role in clinical decision-making.

This study was based on a large clinical cancer database, aiming to explore the potential survival benefit of postoperative adjuvant therapy among EC patients who underwent neoadjuvant treatment and curative surgery.

## Methods

### Research population

All the patients included in this study were sourced from the National Cancer Institute’s Surveillance, Epidemiology, and End Results (SEER) database registry (https://seer.cancer.gov/). The SEER*Stat database, Incidence - SEER Research Plus Data, 17 Registries, Nov 2022 Sub (2000-2020), was utilized as the most recent source of information. Following the acquisition of access permissions, all data were freely accessible in the SEER database. This research was conducted following the principles of the 1964 Helsinki Declaration. As per the determination of our institutional ethics committee, this study is not considered human subjects research and does not require approval from an institutional review board, as it utilizes previously identified data from secondary research sources.

In the SEER database, a total of 77,768 records of esophageal cancer patients were identified, including 39,352 patients diagnosed between 2007 and 2020. Among them, 6,353 patients received neoadjuvant therapy (NCRT, defined as preoperative neoadjuvant radiotherapy or neoadjuvant chemotherapy). Furthermore, we excluded 525 patients with stage IV status esophageal cancer and 23 patients with missing tumor staging information, resulting in a cohort of 5,805 patients for subsequent analysis (Fig 1).

**Fig 1.**
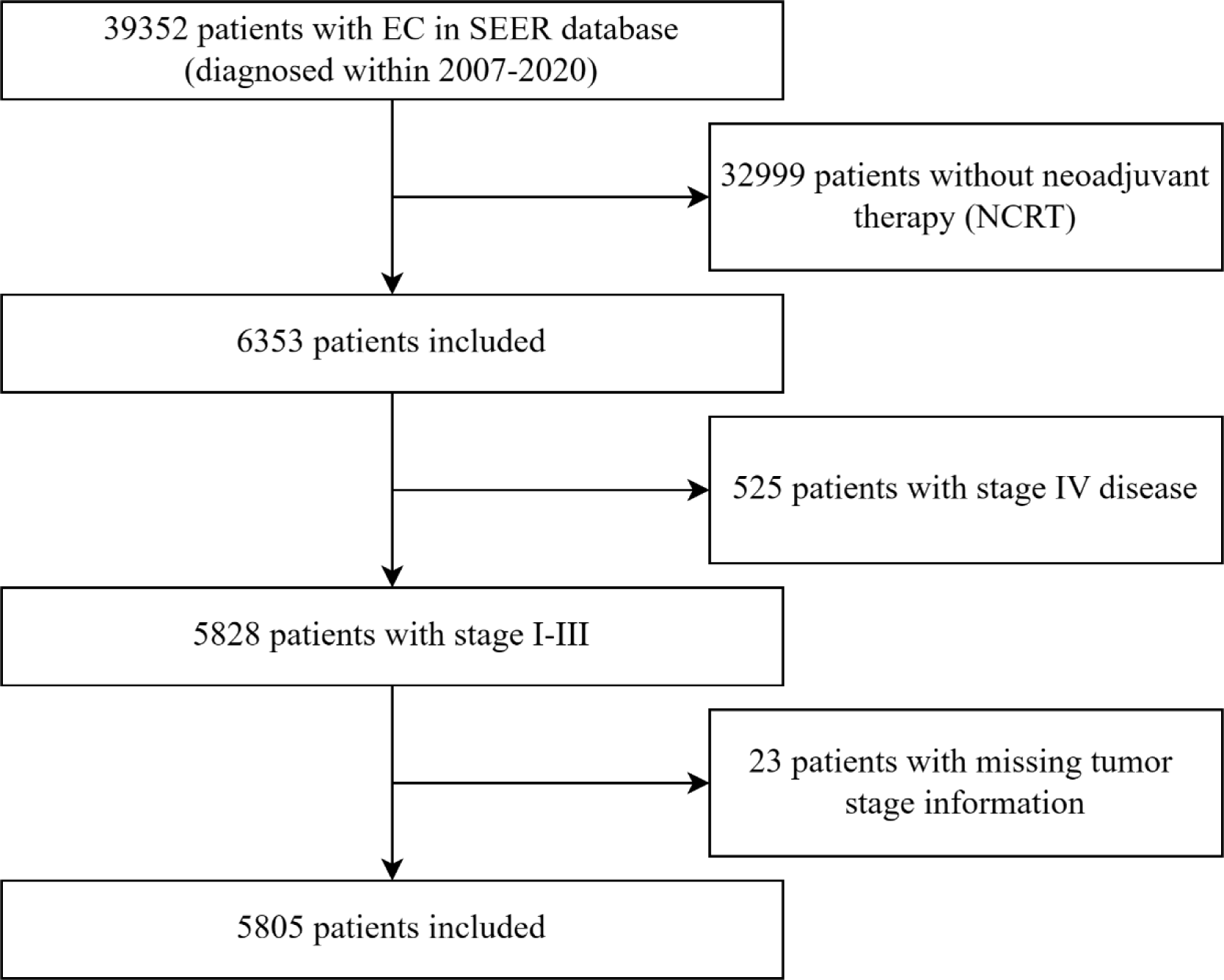
The flow chart of this retrospective cohort study. EC, esophageal cancer; SEER, Surveillance, Epidemiology, and End Results; NCRT, neoadjuvant radiotherapy or neoadjuvant chemotherapy.

We extracted the year of diagnosis, age, race, gender, marital status, tumor staging information, tumor differentiation grade, pathological histology, treatment details, and survival information from the database for the patients included in our study.

### Statistical analysis

Patients were divided into the ACRT group (defined as patients receiving postoperative adjuvant radiotherapy or adjuvant chemotherapy) and the no-ACRT group (without any type of adjuvant therapy), based on whether they underwent postoperative adjuvant therapy. The relationship between the application of ACRT and various clinical variables was examined using the chi-square test or Fisher’s exact test. We employed the propensity score matching (PSM) method to establish the matched cohort for analysis by using the MatchIt package in the R project. PSM algorithm that took all potential confounders into account was conducted at a 1:1 ratio using the nearest-neighbor method with a caliper value of 0.02 to determine matched study cohorts.

The Kaplan-Meier method was employed to depict survival curves, while the log-rank test was used to assess the statistical significance of overall survival (OS) and cancer-specific survival (CSS) differences between the two groups. Univariate Cox regression analysis was conducted to identify predictive factors significantly associated with OS and CSS in the study cohort. The Fine-Gray competing risk model was utilized to control other causes of mortality (OCM) and examine CSS differences between different treatment groups. Multivariate models were applied to determine the impact of ACRT on survival after adjusting for various confounding factors. Additionally, subgroup analyses were conducted to investigate the potential clinical subgroups that may benefit from the treatment. A sensitivity analysis for the EC individuals diagnosed after 2017 was conducted to confirm our results.

P values less than 0.05 were considered statistically significant. All statistical analyses were conducted using R software version 4.2.0 (http://www.r-project.org/).

## Result

### Demographic and clinicopathological characteristics

This study included 5805 patients with histologically confirmed EC based on the inclusion and exclusion criteria (Fig 1). All registered patients underwent neoadjuvant radiotherapy or neoadjuvant chemotherapy (NCRT). Among them, 4838 (83.3%) were male, and 967 (16.7%) were female. There were 3116 individuals (53.7%) over 65 years old in this study population. The study population was predominantly composed of the White race, with 5251 (90.5%) individuals. Among those who received neoadjuvant therapy, 5683 (97.9%) patients received neoadjuvant radiotherapy, while 5741 (98.9%) patients received neoadjuvant chemotherapy, and 5619 (96.8%) patients received neoadjuvant chemoradiotherapy. Regarding postoperative adjuvant treatment, 837 (14.4%) patients received adjuvant chemotherapy or adjuvant radiotherapy (ACRT), with 321 (5.5%) patients undergoing postoperative adjuvant radiotherapy and 672 (11.6%) patients receiving postoperative adjuvant chemotherapy. In terms of the TNM pathological stage, there were 451 (7.8%) cases of stage I, 2178 (37.5%) cases of stage II, and 3176 (54.7%) cases of stage III, respectively.

We grouped patients based on whether they received postoperative adjuvant therapy (ACRT) and compared baseline characteristics between the two groups (Table 1). We found significant associations between postoperative adjuvant therapy application and age (P = 0.010), gender (P = 0.016), N stage (P < 0.001), and TNM stage (P < 0.001). We applied the PSM method to mitigate the influence of confounding factors and balance the baseline characteristics between the two treatment groups. In the end, a total of 1660 EC patients were included in the matched study cohort, with 830 individuals in both the no-ACRT group and the ACRT group. Baseline comparisons and the distribution of propensity scores indicated the two groups were well-matched (Table 1 and Fig S1). The matched cohort was subsequently used as the primary analytical cohort.

**Table 1.**
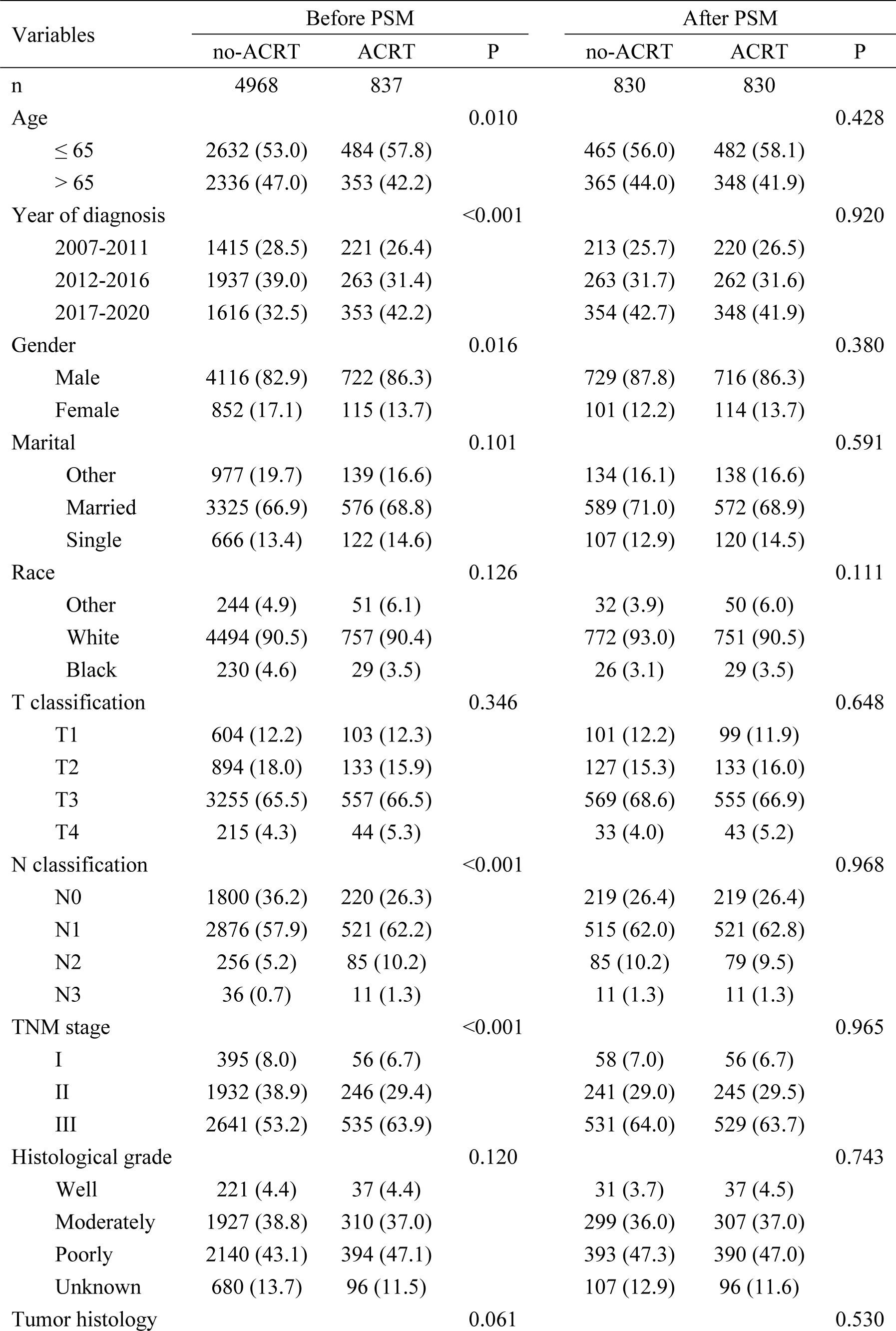
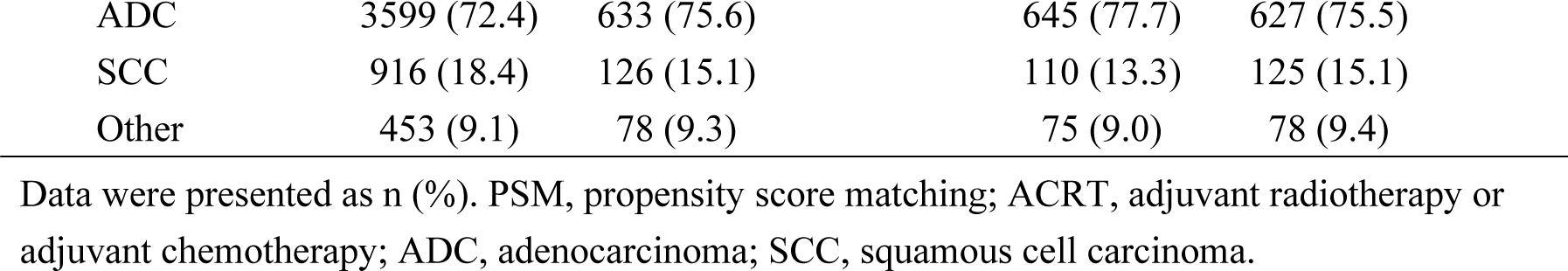
Baseline characteristics of cohorts before and after propensity score matching to examine the difference with or without ACRT after receiving NCRT and surgery.

### Survival analysis

After PSM, the median follow-up duration for the cohort was 59.0 months (95% CI: 54.0-62.0). We compared the survival outcomes of different treatment groups using the Kaplan-Meier method (Fig 2). The median overall survival (OS) in the no-ACRT group was 36.0 months (95% CI: 32.0-43.0), while the ACRT group had a median OS of 34.0 months (95% CI: 30.0-41.0). There was no significant difference for OS between the two groups (P = 0.89). A similar result was observed for CSS: the median CSS in the no-ACRT group and the ACRT group was 49.0 months (95% CI: 40.0-60.0) and 40.0 months (95% CI: 32.0-46.0), respectively, which was without statistically significant difference (P = 0.16).

**Fig 2.**
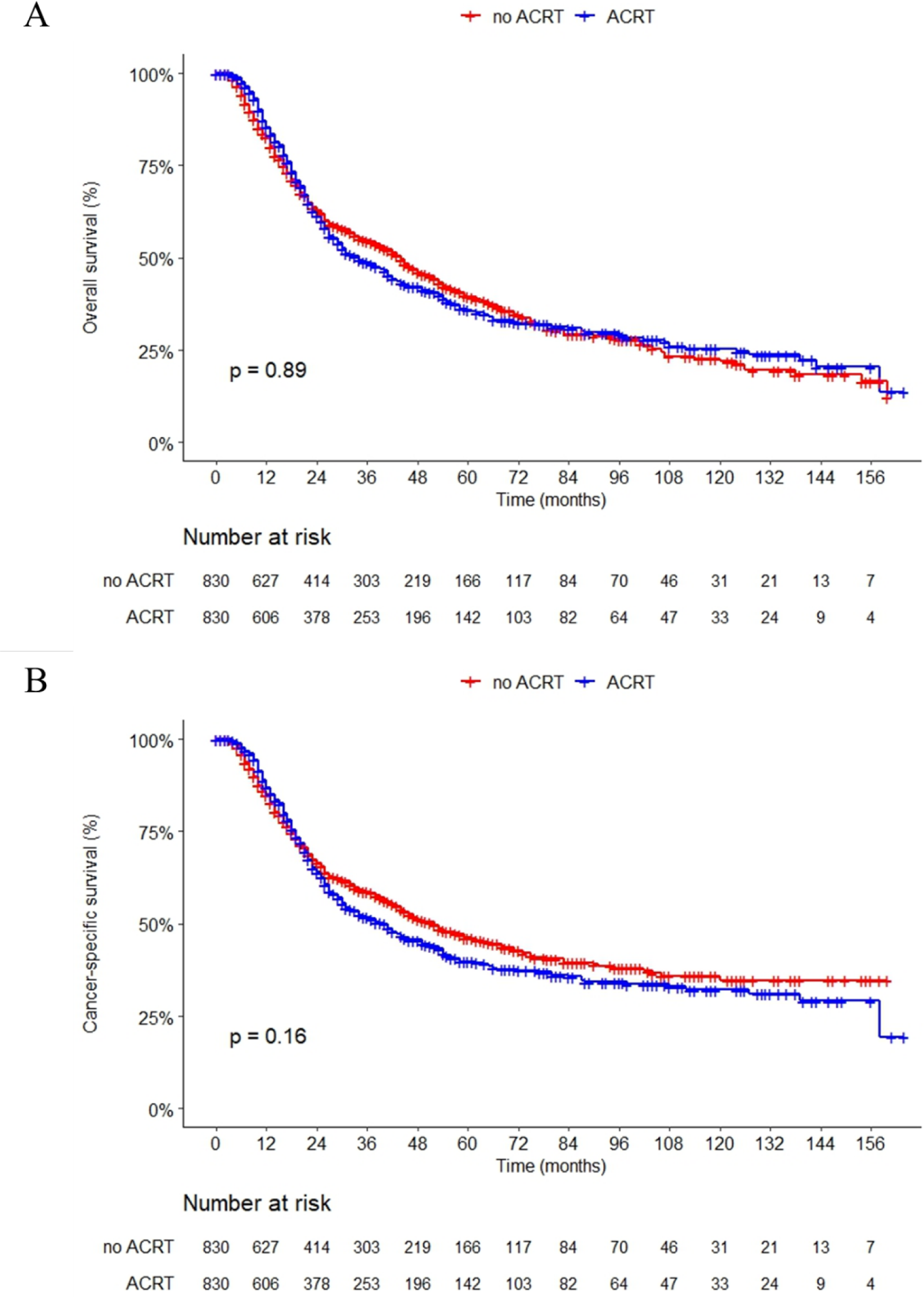
**Overall survival (A) and cancer-specific survival (B) grouped by ACRT in the matched cohort.**

The univariate COX regression model was employed to identify risk factors associated with survival (Table S1). Gender (P = 0.012), T stage (P < 0.001), N stage (P = 0.005), TNM stage (P < 0.001), and tumor histological grade (P < 0.001) were found to be significantly correlated with CSS. However, there was no significant impact of ACRT on CSS (HR = 1.076, 95% CI: 0.932-1.241, P = 0.317) or OS (HR = 0.980, 95% CI: 0.859-1.118, P = 0.763). The multivariate COX regression model did not provide evidence of associations between ACRT and the prognosis of EC patients (Table 2).

**Table 2.** Multivariate COX models and Fine-Gray models for survival analysis.

|  | no-ACRT | COX-CSS |  | COX-OS |  | FGM-CSS |  |
| --- | --- | --- | --- | --- | --- | --- | --- |
|  |  | HR (95% CI) | P | HR (95% CI) | P | HR (95% CI) | P |
| Model 1 | Ref | 1.111 (0.962-1.283) | 0.154 | 1.010 (0.885-1.153) | 0.883 | 1.135 (0.984-1.308) | 0.082 |
| Model 2 | Ref | 1.105 (0.957-1.277) | 0.174 | 1.009 (0.884-1.152) | 0.891 | 1.127 (0.977-1.299) | 0.100 |
| Model 3 | Ref | 1.102 (0.954-1.274) | 0.187 | 1.010 (0.884-1.154) | 0.882 | 1.124 (0.975-1.297) | 0.110 |
| Model 4 | Ref | 1.097 (0.949-1.268) | 0.211 | 1.007 (0.881-1.150) | 0.920 | 1.125 (0.975-1.297) | 0.110 |
| Model 5 | Ref | 1.087 (0.940-1.257) | 0.259 | 0.998 (0.874-1.141) | 0.982 | 1.118 (0.969-1.290) | 0.130 |

Model 1: no adjustment;
Model 2: adjusted for age, gender, year of diagnosis;
Model 3: model 2 + T classification, N classification, TNM stage;
Model 4: model 3 + tumor histology, histological grade;
Model 5: model 4 + race, marital status.
no-ACRT group was set as a reference.
ACRT, adjuvant chemotherapy or radiotherapy; Ref, reference; CSS, cancer-specific survival; OS, overall survival; FGM:
Fine-Gray model; HR, hazard ratio; CI, confidence interval.

We also employed the Fine-Gray competing risk model to assess the impact of postoperative ACRT on CSS (Fig S2), with OCM considered as competing risk events in the model. After adjusting for OCM, the application of postoperative ACRT demonstrated no significant effect on CSS (P = 0.081). The multivariate Fine-Gray model that adjusted for confounding factors similarly did not yield any positive findings (Table 2).

### Subgroup analysis

To assess whether postoperative ACRT might be effective in a particular clinical subgroup, we conducted subgroup analyses using the COX regression model and the Fine-Gray model (Fig 3). In the analysis of CSS, we found that ACRT increased the cancer-specific death risk in the T3 subgroup of EC patients (HR = 1.220, 95% CI: 1.028-1.448, P = 0.023) and in the N1 subgroup (HR = 1.230, 95% CI: 1.031-1.469, P = 0.022). Conversely, in the N2 subgroup (HR = 0.640, 95% CI: 0.382-1.073, P = 0.090) and N3 subgroup (HR = 0.302, 95% CI: 0.112-0.811, P = 0.018), ACRT showed a protective effect, although without a statistical significance in N2 subgroup. The Fine-Gray model yielded consistent results for the T3 (HR = 1.238, 95% CI: 1.046-1.466, P = 0.013), N1 (HR = 1.250, 95% CI: 1.049-1.490, P = 0.013), N2 (HR = 0.636, 95% CI: 0.383-1.057, P = 0.081) and N3 (HR = 0.306, 95% CI: 0.102-0.917, P = 0.034) subgroups. Furthermore, in the subgroups with poorly differentiated tumors, we observed that ACRT had a negative effect on the CSS (HR = 1.243, 95% CI: 1.024-1.510, P = 0.028).

**Fig 3.**
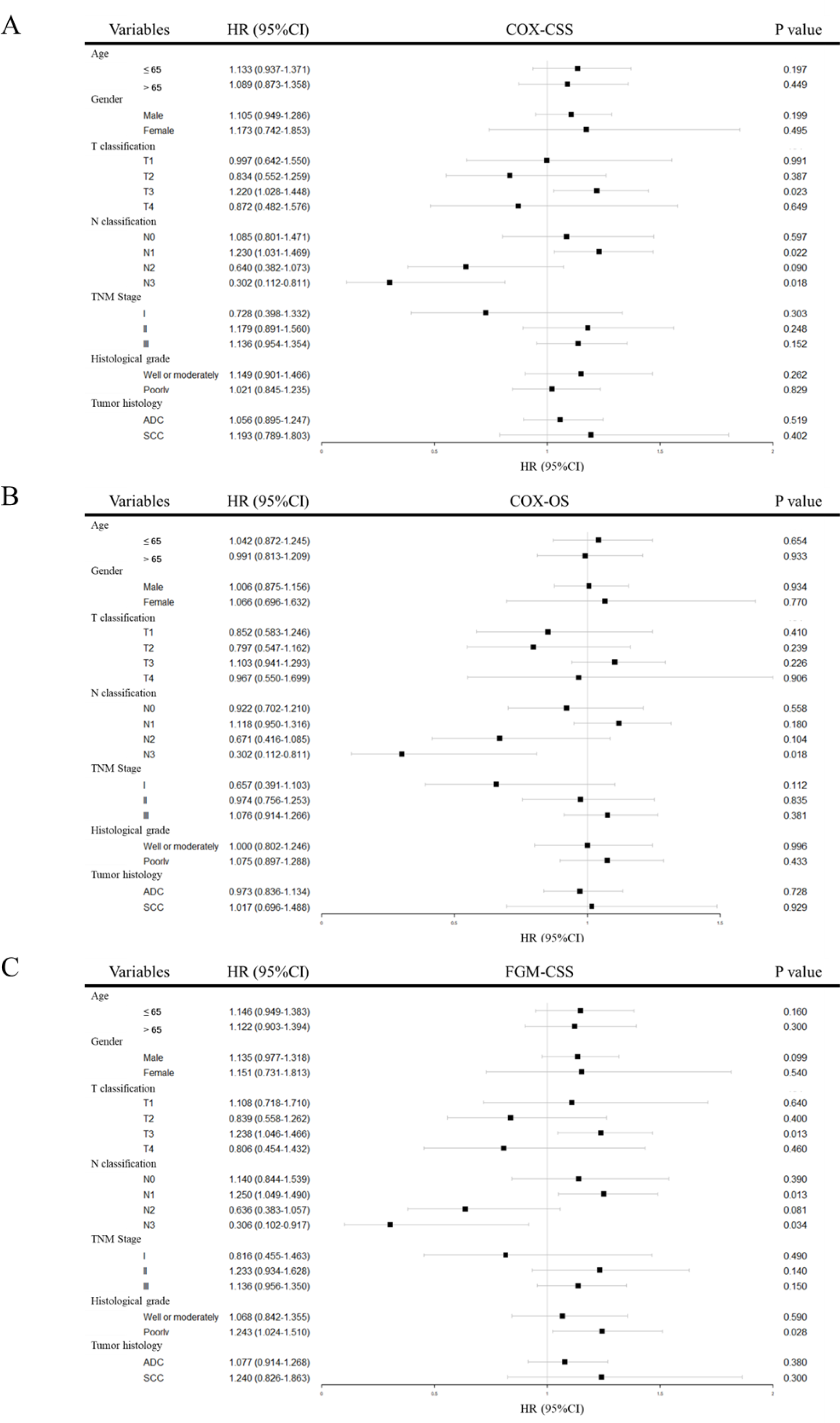
Forest plot of COX model and Fine-Gray model for subgroup analysis. (A) COX model for CSS analysis; (B) COX model for OS analysis; (C) Fine-Gray model for CSS analysis. CSS, cancer-specific survival; OS, overall survival; FGM, Fine-Gray model; ADC, adenocarcinoma; SCC, squamous cell carcinoma; HR, hazard ratio; CI, confidence interval.

### Sensitivity analysis

We conducted a sensitivity analysis to confirm the robustness of our findings (Figure S3). We performed Kaplan-Meier analysis and univariable Fine-Gray regression model analysis on EC patients diagnosed after 2017. The results were consistent with the previous findings, indicating that postoperative ACRT had no significant impact on the long-term prognosis of EC patients.

## Discussion

In this study, we conducted a retrospective analysis of EC patients in the United States SEER database from 2007 to 2020. The results indicated that, overall, among EC patients with neoadjuvant therapy prior to surgery, postoperative adjuvant therapy could not significantly improve survival.

In recent years, there have been varying opinions regarding the role of adjuvant therapy following curative surgery in improving survival. Lee et al. published a meta-analysis in 2022, indicating that the administration of adjuvant therapy after neoadjuvant therapy and surgical treatment could improve the overall survival of esophageal cancer patients, although its impact on disease-free survival (DFS) remained unclear [14]. This finding contradicted our research conclusion. It was worth noting that Lee’s study only included EC patients with negative resection margins while our study enrolled both negative and positive margin statuses due to the lack of margin status information in the database. Additionally, the work of Lee et al. focused on 1-year and 5-year overall survival rates, whereas our study has a longer follow-up period.

In contrast to the findings of Lee et al., a meta-analysis conducted by Malthaner et al. indicated that receiving postoperative adjuvant radiotherapy after preoperative neoadjuvant radiotherapy would result in significantly higher mortality rates [17]. Another retrospective study suggested that postoperative adjuvant radiotherapy following neoadjuvant chemotherapy and surgical treatment for EC patients did not result in improved cancer-specific survival, but even led to worse overall survival [15]. A network meta-analysis of Pasquali et al. revealed that postoperative adjuvant therapy did not offer a survival advantage compared to surgery alone [18]. In our study, we combined adjuvant chemotherapy and adjuvant radiotherapy as adjuvant therapy (ACRT) for analysis, and the results demonstrated that ACRT did not substantially improve OS or CSS in patients who underwent NCRT and surgery treatment. Although our approach of defining neoadjuvant chemotherapy and neoadjuvant radiotherapy together as NCRT may introduce bias to a certain extent, previous research has not identified survival differences between neoadjuvant chemotherapy and neoadjuvant radiotherapy [19, 20], suggesting that our classification method may be appropriate.

In general, there is a tendency to administer additional adjuvant therapies to postoperative patients who are found to be in more advanced stages, including those with a greater number of lymph nodes involved and margin-positive resection of EC patients [8, 21, 22]. In the exploratory subgroup analysis of clinical cohorts in our study, ACRT demonstrated a protective effect on the prognosis of EC patients in the N2 and N3 subgroups, which was consistent with the findings of Matsuura et al. [23]. However, due to limitation of subgroup sample size, caution should still be exercised in interpreting this result. Some studies have suggested that EC patients with lymph node positive can benefit from adjuvant chemotherapy [8, 24], but our subgroup analysis did not support a survival benefit from adjuvant therapy in this subgroup of patients (HR = 1.134, 95% CI: 0.965-1.334, P = 0.130). This may imply that ACRT exhibits a survival advantage only in EC patients with a higher burden of lymph node metastasis. In the N1 subgroup, ACRT even increased the risk of cancer-specific death, and additionally, it had a negative impact on survival in T3 status and poorly differentiated EC patients, which might be explained by potential confounding factors, such as surgical margin status, as patients with positive margins are more likely to receive postoperative adjuvant therapy. In contrast to previous studies [25], our subgroup analysis did not suggest an advantage of ACRT for squamous cell carcinoma.

In the sensitivity analysis, we utilized the most recently released SEER data (EC patients diagnosed after 2017), which led us to believe that the most appropriate therapy regimens were used for the corresponding patients, thus mitigating the confounding factors arising from various therapy regimens to a certain extent. Similar results corroborated the reliability of our conclusion, albeit using a relatively smaller sample size and a shorter follow-up duration.

Based on the results of this study, we should contemplate whether it is advisable to recommend postoperative adjuvant therapy for esophageal cancer patients, as it may not improve overall survival. Postoperative adjuvant therapies inevitably result in toxicity side effects and economic burdens [26–28]. Research indicated that the tolerability of adjuvant therapy was significantly worse compared to neoadjuvant therapy [29]. Esophageal cancer patients undergoing radiation therapy are more susceptible to developing thoracic tumors [30]. Furthermore, studies have shown that postoperative adjuvant radiotherapy had a significant association with severe cardiovascular events and pulmonary function changes [31, 32]. These findings underscore the necessity of personalized treatment for patients, requiring more meticulous efforts to identify the EC patients who would benefit from postoperative adjuvant therapy. Patient selection for adjuvant therapy based on omics data may be a promising solution [33].

In recent years, researchers have been exploring immunotherapeutic approaches for esophageal cancer, aiming to provide new treatment options other than postoperative adjuvant radiotherapy and chemotherapy [34]. The CheckMate 577 trial demonstrated that, when compared to a placebo, the application of postoperative immunotherapy in EC patients who have undergone neoadjuvant therapy and curative surgery could significantly improve the DFS of EC patients [35]. Furthermore, several studies related to immunotherapy are currently underway [36], and we look forward to the publication of their results.

Our study has certain limitations: Firstly, its retrospective nature and the absence of some data may introduce bias in data interpretation. The lack of surgical margin information could significantly impact the study results, as it is a crucial confounding factor in the research. Secondly, there is no information regarding the therapy regimen, radiation dose, treatment duration, and adverse events, making it impossible to compare the efficacy of different therapy regimens. Furthermore, the data for this study exclusively comes from Western populations, and therefore, cannot be extrapolated to other ethnic groups.

## Conclusion

After neoadjuvant therapy and curative surgery treatment, the application of adjuvant therapy in esophageal cancer patients has shown limited survival benefits, and further prospective research is required to validate this observation. Only in EC patients with a more advanced N status, the significant survival benefit was observed. It is imperative to explore more suitable postoperative adjuvant therapy strategies to alleviate the health and economic burdens on esophageal cancer patients.

## Data availability statement

All the data used in this study were publicly available in the SEER database (https://seer.cancer.gov/).

## Abbreviations

ACRT: adjuvant radiotherapy or adjuvant chemotherapy
ADC: adenocarcinoma
CI: confidence interval
CSS: confidence interval
DFS: disease-free survival
EC: esophageal cancer
FGM: Fine-Gray model
HR: hazard ratio
NCRT: neoadjuvant radiotherapy or neoadjuvant chemotherapy
OCM: other cause mortality
OS: overall survival
PSM: propensity score matching
SCC: squamous cell carcinoma
SEER: Surveillance, Epidemiology, and End Results.

## Author contributions

Weiyi Jia and Renwang Hu conceived the study design. Weiyi Ji and Chao Li were involved in data analysis. Weiyi Jia and Can Liu were involved in the figure preparation. Weiyi Jia was the major contributor to writing the manuscript. All authors read and approved the final manuscript. The work reported in the paper has been performed by the authors, unless clearly specified in the text.

## Acknowledgement

Not applicable.

